# Confronting COVID-19: Surging critical care capacity in Italy

**DOI:** 10.1101/2020.04.01.20050237

**Authors:** Jose M Rodriguez-Llanes, Rafael Castro Delgado, Morten Gram Pedersen, Pedro Arcos González, Matteo Meneghini

**Author notes:** three are last authors (Morten Gram Pedersen, Pedro Arcos González, Matteo Meneghini).

## Abstract

The current spread of severe acute respiratory syndrome coronavirus 2 (SARS-CoV-2) in Europe threats Italian’s capacity and that of other national health systems to effectively respond to the needs of patients who require intensive care, mostly due to pneumonia and derived complications from concomitant disease and age. Predicting the surge in capacity has proved difficult due to the requirement of a subtle combination of diverse expertise and difficult choices to be made on selecting robust measures of critical care utilization, and parsimonious epidemic modelling which account for changing government measures. We modelled the required surge capacity of ICU beds in Italy exclusively for COVID-19 patients at epidemic peak. Because new measures were imposed by the Italian government, suspending nearly all non-essential sectors of the economy, we included the potential impacts of these new measures. The modelling considered those hospitalized and home isolated as quarantined, mimicking conditions on the ground. The percentage of patients in intensive care (out of the daily active confirmed cases) required for our calculations were chosen based on clinical relevance and robustness, and this number was consistently on average 9·9% from February 24 to March 6, 2020. Five different scenarios were produced (two positive and three negative). Under most positive scenarios, in which R_0_ is reduced below 1 (i.e., 0 ·71), the number of daily active confirmed cases will peak at nearly 89 000 by the early days of April and the total number of intensive care beds exclusively dedicated to COVID-19 patients required in Italy estimated at 8791. Worst scenarios produce unmanageable numbers. Our results suggest that the decisive moment for Italy has come. Jointly reinforcement by the government of the measures approved so far, including home confinement, but even more important the full commitment of the civil society in respecting home confinement, social distancing and hygiene will be key in the next days. Yet, even under the best circumstances, intensive care capacity will need to get closer to 9000 units in the country to avoid preventable mortality. So far, only strong measures were effective in Italy, as shown by our modelling, and this may offer an opportunity to European countries to accelerate their interventions.

## Introduction

The epicenter of the global COVID-19 pandemic seems to have relocated to Europe while fast spread continues worldwide (1-3). While mathematical models and ground experience consistently show that strong government response measures did work in China to slow down the spread of SARS-CoV2 (4, 5), there is now a pressing need for a coordinated upgrade of health systems in Europe to cope with this unprecedent crisis while government interventions come into place. The World Health Organization reports fast growing epidemic curves in Iran, Italy, Spain, France and Germany (1,3). The Achilles’ heel of any health system is the limited capacity to deal with high patient inflows in a short period of time, and particularly patients that use scarce resources such as intensive care units (ICU) or mechanical ventilation (6-10).

This crisis combines both characteristics (figure 1). Patients raise exponentially in numbers and a significant share of those patients will develop severe pneumonia (10), aggravated by concomitant disease and age (11, 12). In addition, health workers are heavily exposed and they are overrepresented amongst the sick (13, 14), which undermines the response capacity of the health system. The consolidated experience from China in responding to this crisis (5) and the lessons learnt from Italy in the early phase of the epidemic have the potential to provide important guidance to other nations. The two experiences provide a clear message that this epidemic can overwhelm health care capacities in a matter of days (5, 15, 16). While the Italian government reacted quickly to early cases detected in Codogno by declaring the State of Emergency, followed by progressively restrictive measures (Box), the epidemic is not yet under control, with new cases ranging from 3 to 5 thousands per day in the last few days (17).

**Figure 1.**
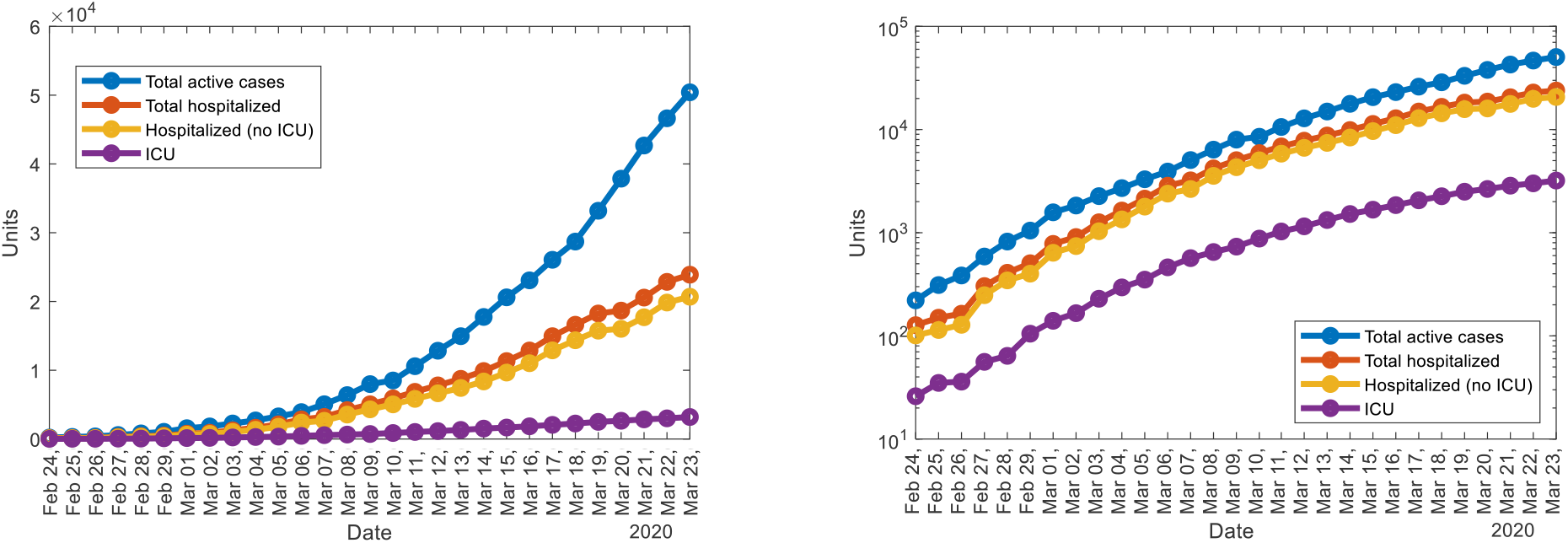
Daily active cases of SARS-CoV2 total active cases, total hospitalized patients, hospitalized patients (not in ICU), ICU patients in Italy, Feb 24-Mar 23, 2020. **A**. Left: linear vertical scale. **B** Right: logarithmic vertical scale.

Estimating national surge capacity for critical care utilization (i.e., ICU bed) is possible and different approaches have been recently reported (15, 16). While they timely raised awareness and public concern on the desperate need to raise the number of intensive care units in Italy, their predictions proved difficult (15, 16). By analyzing the epidemiological curve carefully considering government interventions, it is possible to estimate the number of critically ill patients and adapt surge capacity before the system is overwhelmed.

**Box**. Milestones of government interventions regarding COVID-19 epidemic in Italy

| Date | Intervention name (societal implications) (18) |
| --- | --- |
| January 31, 2020 | Declaration of State of Emergency |
| February 23, 2020* | Containment measures in a Red Zone of 11 municipalities in North Italy (lockdown, social gatherings suspended, social distancing, educational services closed and only allowed remote work and productive activities requiring physical presence) |
| March 1, 2020 | Containment measures in Red Zone now expanded to 3 regions and 2 provinces in North Italy (lockdown, social gatherings suspended, social distancing, educational services closed and only allowed remote work and productive activities requiring physical presence) |
| March 8-11, 2020 | Expansion of Red Zone to the entire country (Mandatory home confinement of the entire country. Productive activities requiring physical presence allowed nationwide). |
| March 22, 2020 | In addition to the above. All productive activities requiring physical presence nationwide are paused except essential ones (only those related to food and health). |

*Schools closings started in some regions of the Northern Italy, including Lombardy and Veneto.

We argue that addressing this challenge requires a combination of diverse disciplines including meaningful epidemiological measures, concepts on critical care resource utilization, outbreak modelling and some dose of pragmatism. We estimated the maximum number of required ICU beds exclusively dedicated to COVID-19 patients, that will be required at the peak of the epidemic, while considering previous impact of government measures and five possible scenarios by new restrictions implemented on 22 March 2020.

### Estimating critical care utilization: current approaches

We identified two types of approaches. The first set combines clinical data, such as percentage of COVID-19 patients using an ICU bed before study cutoff (16) or epidemiological data, i.e. daily numbers of occupied critical care beds out of active laboratory confirmed cases (15), supplemented with exponential prediction models (15, 16). In one report, the number of patients who were actively infected were fitted to an exponential model. The same model exponent was used to fit the number of patients in intensive care units. In another report, a linear and exponential models were fitted to the number of ICU bed admissions expected. Importantly, none of the predictions made did match ground data few days later (15, 16).

An obvious limitation of both approaches is its transitory use, given that the epidemic is not expected to raise infinitely neither move following a linear trend. This shows that prediction itself is not that straightforward in this context. Moreover there are implicit dynamics in the flow of infected, sick, recovered and deaths which affect the flow of inpatients and the available ICU beds. In that context a model of the SIR family can account for these interconnected quantities in a dynamic framework (19).

A second limitation is that they cannot easily accommodate the government measures and their impact on the evolution of the disease. In Italy, for example, early containment measures (school closings, hygiene indications) were followed by more restrictive regional lockdown and social distancing measures. By March 11, a far more restrictive national lockdown was declared. On March 21, the Italian Prime Minister Giuseppe Conte communicated that the non-essential productive activities would be suspended within the country in an attempt to decelerate even more the epidemic ascension (box).

A third limitation observed in one of the studies in particular (16) is the baseline numbers used. A 5% of ICU bed requirement from China (10) is far too low for various reasons. Most hospitalized patients were still hospitalized at the time of study cutoff, meaning that probably this 5% underestimated the true UCI bed requirements for the entire length of stay of all patients in that sample. Moreover, that was the proportion of a sample of hospitalized confirmed patients and not of the confirmed active cases. The largest study from the Chinese CDC did not offer a better number, despite being a sample of confirmed cases, they provided a proportion of critical patients (4·7%) certainly requiring intensive care, but a larger proportion of severe cases (13·9%), which under certain conditions may also require an ICU bed (13). Crucially, none of these numbers with one exception (15) consider the dynamic nature of the rate of use of ICU beds. In that study, the percentage of patients in intensive care reported daily in Italy between March 1 and March 11, 2020, has consistently been between 9% and 11% of patients who were actively infected (15).

The second set of approaches is promoted by SIRS models, which scientists feed with the available information on the characteristics of the epidemic, disease course and epidemiology (for example numbers of infected and seeking care, hospitalization bed rates, UCI admission rates) and government and civic measures adopted. The main limitation of these models is that they rely on a sequence of equations based on best available numbers (20). As shown earlier, some of these numbers proved to be very incorrect and can have a vast impact on calculations and predictions. While we applaud these efforts to provide a range of policy options and their consequences on health and demand for critical care, such an undertake also includes large uncertainty in the numbers produced. The authors reported that certain policy options were robust to model uncertainty derived from potential variability in essential parameters such as the severity of the virus as captured by the proportion of cases requiring ICU admission or the basic reproduction number R_0_ (20). Other studies show how vast these uncertainties can be, for the mere prediction of the course of this epidemic (21).

### EpiCare approach

We proposed a hybrid approach, called here EpiCare, that gives equal relevance to the modelling approach and the epidemiological measures on health care used to estimate surged critical care capacity. The modelling approach itself is dominated by two principles: on one hand, parsimony and selection of a limited number of robust parameters to reduce uncertainty. On the other hand, the model requires previous validation with ground data (i.e., confirmed active cases, hospitalization rates) and additional testing of its predictive ability in a 5-10 day window to validate its usefulness for planning in at least the following 10 days. Consequently, especial attention is given to the choice of measures and their robustness over time.

We defined our primary outcome as the national Daily ICU rate, expressed as the percentage of daily patients in intensive care units in Italy out of persons who were actively infected and reported daily in Italy (17). All patients in numerator and denominator were SARS-CoV2 laboratory confirmed cases. We did not recommend using alternative denominators, such as the total cumulative number of confirmed cases, as the percentage need to reflect the probability of a confirmed active case to be in an ICU on a given day.

We then examined the robustness of this measure over time using provided data by the Italian Civil Protection (17). Additionally, we investigated by means of visual plots the national rate of daily increments in ICU patients by the daily increment in active confirmed cases, expressed as a percentage (Figure 2 A). Figure 2 B shows early surging of ICU patients on the first days of the epidemic 24-25 February, and a second surge in ICU use peaking around 6 March. Additional regional analyses (not shown) confirmed that case surges in neighboring regions of Lombardy such as Piemonte and Liguria were contributors for the national rise in ICU cases 10 days later. From March 12, a steady decrease is noted and this is further accentuated after March 18 (Figure 2 B). As previously reported (15) these rates were mostly within the range of 9% and 12% of active confirmed cases. Based on these observations, we delineated four relevant periods (table 1). We based our choice of period to calculate the Daily ICU rate on three aspects. First, the two early periods up to 6 March are likely more representative of ICU admission criteria based on clinical realities. Secondly, a higher value will support a more conservative estimate of ICU bed requirement (9.9%). Third, high value consistency across these the first two periods.

**Table 1.** Average rate of ICU beds used per active SARS-CoV2 confirmed cases for four periods in Italy, 24 Feb-21 March, 2020

| Critical care periods | Daily ICU rate, mean |
| --- | --- |
| Early phase and case surge in Lombardy, 24-28 Feb | 9.9% |
| Early phase and case surge from neighboring regions, 29 Feb-6 March | 9.9% |
| Surge critical care capacity I, 7-18 March | 9.4% |
| Surge critical care capacity II, 19-21 March | 7.3% |

**Figure 2.**
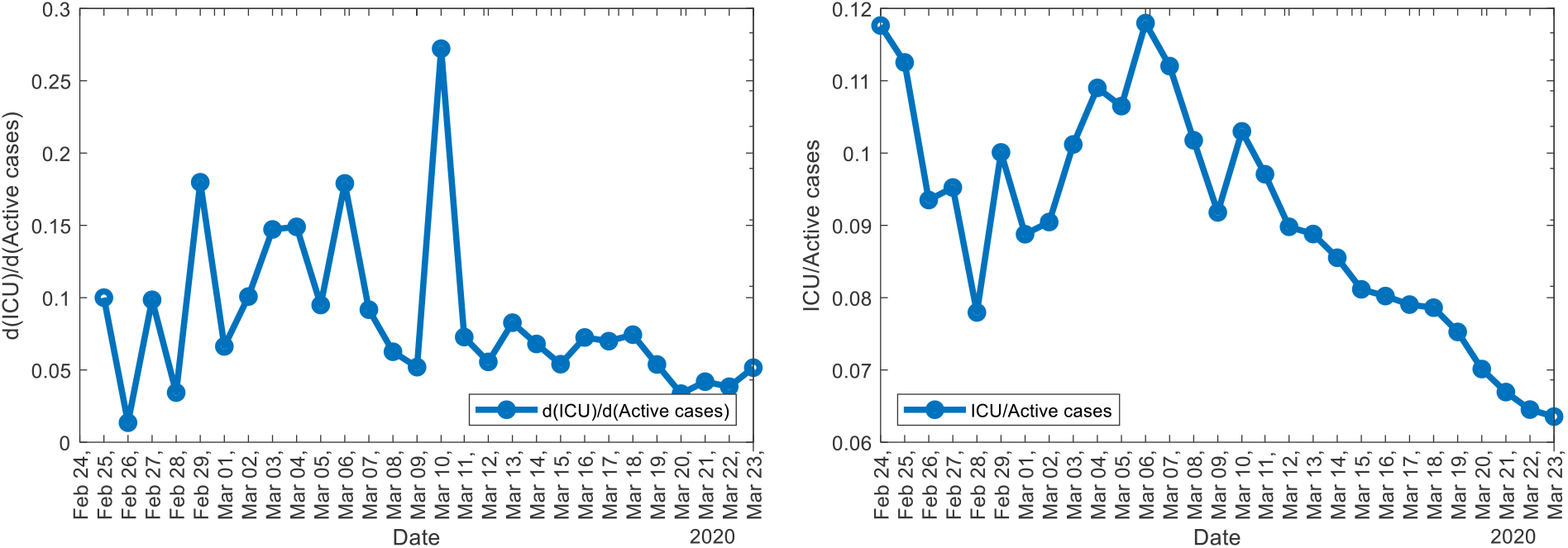
**A** (left) Daily increase of ICU patients divided by daily increase of active cases. **B** (right) Daily rate of ICU beds used per active SARS-CoV2 confirmed cases in Italy, till March 23, 2020

Subsequently we focused on the modelling strategy to forecast the expected number of active cases. Specifically, we decided not to model the spread of the disease by the conventional SIR (susceptible, infectious, recovered) model, since Italy was the first Western country to adopt immediate quarantine to contrast the diffusion of the virus. For this, we used a SIQR model (figure 3), where we consider a sub-population of quarantined individuals (Q). Details on the model and on the related mathematical framework can be found elsewhere (19). The quarantined patients, once identified, are immediately isolated (either in hospital or at home), thus no longer transmitting the disease. It is worth noticing that we decided not to adopt a SEIR approach (susceptible, infectious, exposed and recovered), since it is well known that undetected asymptomatic individuals can transmit the disease (19). The model was fitted with data on total confirmed active COVID-19 cases up to 19 March, and its predictive capability was checked with the available data up to 24 March. Results were found to be within the 50 % confidence band (indicating less than 50% variation in R_0_) in the subsequent 5-days period, indicating that the model works reasonably for planning resources on the mid-term (figure 4). Longer-term predictions make little sense in a period when new drastic measurements are introduced almost daily (the latest was introduced on March 22). On the other hand, thanks to its intrinsic robustness, our model can be used to compare long-term scenarios, thus allowing to visualize projections on the potential effectiveness of the latest measures implemented.

**Figure 3.**
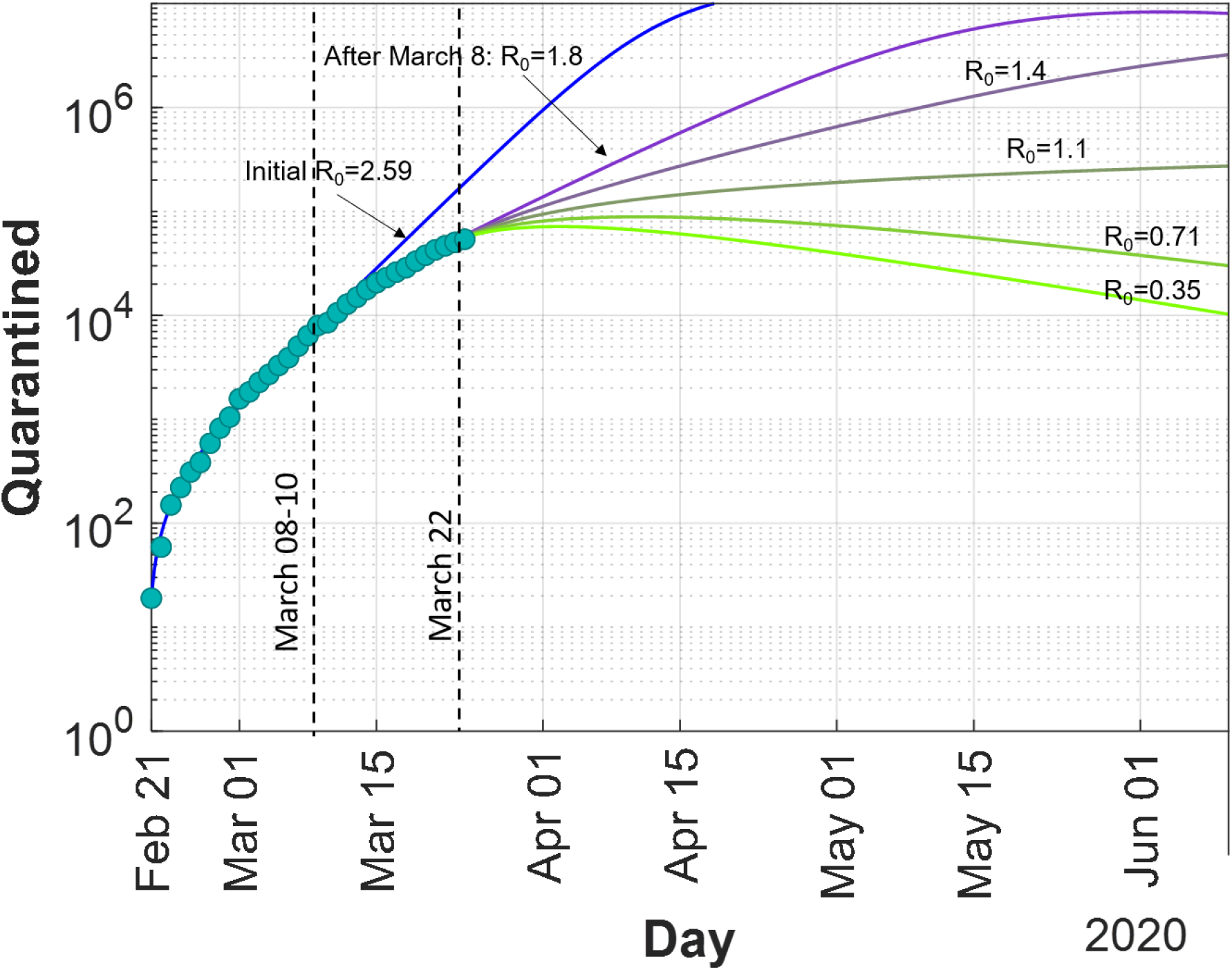
Model predictions of active confirmed SARS-CoV2 cases (home isolated or hospitalized) under four scenarios of progression. Light blue dots are the observed active confirmed cases of SARS-CoV2 daily reported in Italy. Major interventions with impact on slowing down the epidemic curve (8-10 March 2020, R0 from 2·59 to 1·8) and last intervention by 22 March 2020, with a foreseen impact and four possible scenarios of progression. R0 is the reproductive number.

**Figure 4.**
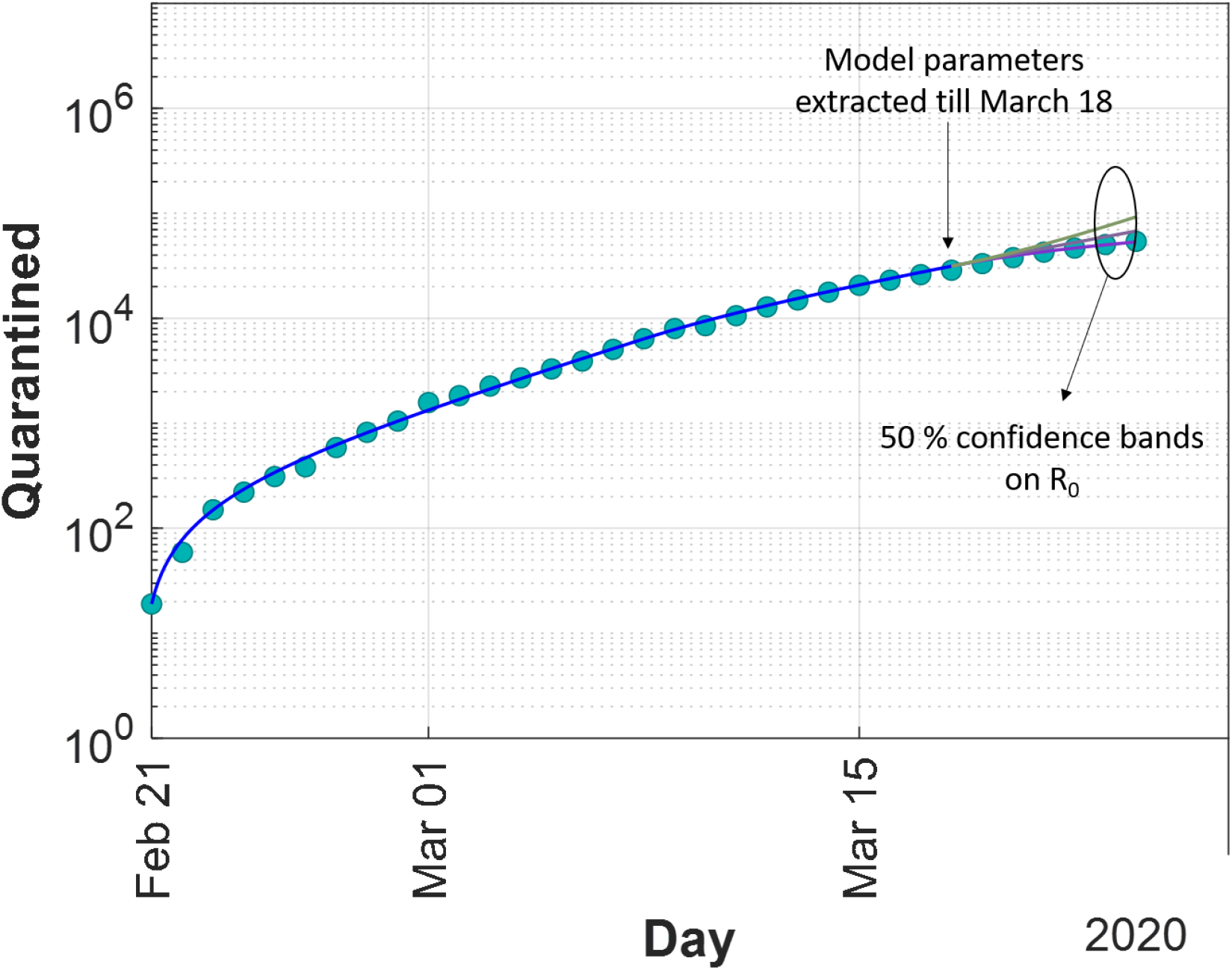
Test of predictive capability of the model

The main parameters of the model were obtained by analytically fitting the total number of cases as recorded by the Italian Civil Protection, and by considerations based on the literature regarding incubation time and disease duration (19). From such parameters, we could estimate the basic reproduction number before the first Italian cases appeared (R_0_=3·5), and after COVID-19 testing and quarantine was introduced (R_0_=2·6). In addition, we estimated the effects of the measures of social distancing introduced after March 8 followed by further extension to the national territory a couple of days later (box 1). Such drastic measures resulted in a decrease of R_0_ from 2·6 to 1·8.

Finally, based on the same set of parameters, we were able to predict the evolution after March 22 (day when further measures were introduced, including closure of most factories and prohibition of moving out of the municipality of residence), based on five different scenarios. Such scenarios consider: R_0_=1·8 (equal to what obtained after the measures introduced on March 08, i.e., no further effect due to the limitations introduced on March 22), R_0_=1·4 (moderate decrease in R_0_), R_0_=1·1 (40 % decrease in R_0_), R_0_=0·71 (similar to the value obtained by the Chinese government in Hubei after restrictions, (19)), R0=0·35 (immediate stop of the disease). Thus, under these assumptions, we were able to estimate the increase of number of cases till May 01. The results clearly indicate that to keep the number of quarantined (sick home isolated and hospitalized patients) people below 90 000, a R_0_ significantly lower than 1 is needed (0.71). Under the assumption that 9·9% is a robust measure of the proportion of those cases that may require ICU treatment on a given day, (even in this scenario) this would mean that the Italian health system would need at least 8791 ICU beds entirely dedicated to COVID-19, as early as 10 April 2020, and till May 1.

## Discussion

We estimated, by using a SIQR model combined with robust daily ICU rates, the surge capacity of critical care (i.e., ICU beds) required in Italy exclusively for COVID-19 patients. In doing so, we considered five different scenarios (two positive and three negative) after last measures were enforced by the government on 22 March 2020.

Under one plausible positive scenarios (similar to the Chinese case, so ambitious), in which R_0_ is pushed substantially below 1 (0·71), the number of daily active confirmed cases will peak at 88 800 by 10-11 of April and the total number of intensive care beds exclusively dedicated to COVID-19 patients would be then estimated at 8791 beds for the entire country.

Under the two other scenarios, more pessimistic but possible, R_0_ higher than 1 promote a later and higher reach of the epidemic peak, which gets out of reach by our predictions. Under these conditions the progression of cases becomes untenable.

Therefore, under such circumstances the number of ICU beds require to provide sufficient care to the severe patients would be unreachable.

One of the critical points in dealing with a surge of severe patients in an epidemic emergency is the constrained capacity to provide critical care (22). In epidemics such as the one the world has to confront now, high reproductive numbers may generate high loads of temporally concentrated severe cases (23).

This has proved to be the case here. From the beginning of this emergency in Italy, the ICU bed capacity was identified as a major bottleneck and strong efforts, commensurate resources and attention have been dedicated to this direction from government, planners, media and academics. These efforts are ongoing. Italy has managed to increase its initial capacity of approximately 5200 ICU beds. Assuming that half of these beds can be exclusively used for COVID-19 patients, around 2600 ICU beds were available at baseline. As of 23 March, 3204 ICU beds were under use for these patients in Italy (17), neatly above baseline capacity under those assumptions. There is evidence that ICU capacity in Lombardy was approximately 720 beds pre crisis (16) while 1183 were used on 23 March (17). The most difficult situation is described in Lombardy where critical care capacity has reached, according to regional government, its maximum capacity (24). There are better news in other regions which are currently increasing ICU capacity such as Veneto (n=325), Toscana (n=745), Campania (n=490) or Piemonte (n=160). Others did not report. These four Italian regions could be adding approximately 1720 new beds. This number is quite similar to numbers collected by the media, 1850 (25). Currently, we estimate that the ICU capacity for this emergency is approximately 5000 beds. According to our data and estimates, under a positive scenario, this number should be increased by 76% progressively in the next two weeks if hospitals are to provide good quality of care (15).

A previous report warned that estimating the peak of the epidemic (15) was the cornerstone of adequate planning in surging the critical care capacity in Italy though difficult to estimate. We hope to have added some light to this pressing question.

Our study is not exempt of limitations. Our rate of daily ICU use is obviously affected by conditions such as changes in case definitions, testing capacity, changes in clinical early detection, and changes in disease severity. We were conscious that case definition and testing strategies to optimize resource use were redirected before the end of February in Italy. Most severe patients probably arrived first to hospitals and a majority of them were of advanced age and with varying levels of comorbidities (11, 14). Equally important, ICU admission criteria may have changed over the course of the emergency to deal with scarce ICU resources in particular locations, with difficult ethical choices to be made between providing ICU treatment versus palliative care in certain patients (26). To counteract these effects we chose a longer period to estimate our daily ICU rates and we corroborated the consistency of these values across days and periods defined. Additionally we chose the period at the beginning of the emergency where decisions were more likely made based on clinical realities and not on allocation of scarce resources. Our models, to a certain extent, depends on the quality of parametrization and its ability to predict future evolution of the epidemic. With this issue in mind, we tested its predictive ability on a six day window and we hope that this is enough to provide a range of good predictions from 25 March on and for at least 10 days, in a context where new drastic measures are introduced almost daily. We are aware that the numbers used in the model parametrization come with their own uncertainty, given that this disease is new, and context relevant data is not always available.

We are also aware that we did not address important issues like the need of assisted ventilation, but we found that the literature is not yet clear, prospective studies are lacking or we were not able to deal with the mounting level of evidence which is being published at extraordinary pace on these days. Of course, the extra resources required to deal with this crisis will require commensurate medical staff to the extra needs pointed out by our study. While we modelled data at Italian level, regional level models would be a natural step to take, if not timely in Italy, at least for other countries in which autonomous regions and regions have to deal with the administration of health care resources. The early lessons learned from Italy should be taken as an opportunity by neighboring countries to learn and act immediately.

## Conclusion

Current provisions of critical care in Italy are not sufficient to deal with the peak of the epidemic that is coming. There is evidence that critical care has been an issue for some time now, particularly in certain regions such as Lombardy where such capacity is not expanding further or at a visible rate (15, 16). Our findings suggest that the situation may deteriorate further and call to a double line of action by government, business sector and civil society (including NGOs) as a whole to act upon this crisis.

First, these above actors may mobilize enough funds for quick and further increase of ICU beds in regions most in need. It seems that regions are not able to easily transfer patients to regions with less ICU patients and where these beds may be available (24). To avoid further and irreconcilable ethical dilemmas to arise in the next days and weeks we must address these issues now and further ICU capacity and transferability of patients should be actioned.

Second, and more than ever, awareness of civil society and the business sector to respect home confinement and limit industrial and productive activity is crucial. Promotion of these positive behaviors should be promoted by government, business sector and citizens through social media, internet, radio and television.

Finally, Italy remains a guiding example for this crisis, now approaching like a tsunami to the rest of Europe. The Italian government quickly reacted and declared the state of emergency after few cases appeared in Codogno. They rapidly activated the coordination of institutions such as the Instituto Superiore di Sanitá and Civil Protection Agency which have been working hand in hand and communicating and coordinating daily to the general public, with enormous transparency, diligence and respect to the victims. In addition, the Civil Protection Agency implemented a modern open data policy regarding epidemiological and health care data in near-real time and with clear definitions and support. This is facilitating a sense of unity, respect and also a real time response of the scientific community.

### Contributors

JMR-L had the research idea, wrote the paper, was responsible for data analysis and statistics, interpretation, and led the work. RCD contributed to analysis and literature, discussion and interpretation of the clinical data in the manuscript. MGP developed the model and contributed to writing of the manuscript. PAG contributed literature, discussion and interpretation of the clinical and epidemiological data in the manuscript. MM was responsible for simulations, data analysis and figures, and contributed to writing of the manuscript. All authors critically revised the manuscript for intellectual content and approved the manuscript before submission. JMR-L, MGP and MM had access to the datasets used and are responsible for the paper.

## Data Availability

All data used in this article is public and open access, free of charge.

https://github.com/pcm-dpc/COVID-19

## Declarations of interest

We declare no competing interests.

### Acknowledgments

We are grateful to Dr. Maryline Kolb for critical discussion on the paper. Activity at University of Padova was partly supported by MIUR (Italian Minister for Education) under the initiative “Departments of Excellence” (Law 232/2016).

## Other declarations

JMR-L works for the European Commission as researcher and he declares that the views expressed are purely those of the writer and may not in any circumstances be regarded as stating an official position of the European Commission. His work is motivated by the duty of care.

